# Resolution of *SLC6A1* variable expressivity in a multi-generational family using deep clinical phenotyping and *Drosophila* models

**DOI:** 10.1101/2024.09.27.24314092

**Authors:** Kristy L. Jay, Nikhita Gogate, Kim Ezell, Jonathan C. Andrews, Sharayu V. Jangam, Paige I. Hall, Hongling Pan, Kelvin Pham, Ryan German, Vanessa Gomez, Emily Jellinek-Russo, Eric Storch, Brain Gene Registry Consortium, Undiagnosed Diseases Network, Shinya Yamamoto, Oguz Kanca, Hugo J. Bellen, Herman Dierick, Joy D. Cogan, John A. Phillips, Rizwan Hamid, Thomas Cassini, Lynette Rives, Jennifer E. Posey, Michael F. Wangler

## Abstract

**Purpose:** Variants in *SLC6A1* result in a rare neurodevelopmental disorder characterized by a variable clinical presentation of symptoms including developmental delay, epilepsy, motor dysfunction, and autism spectrum disorder. *SLC6A1* haploinsufficiency has been confirmed as the predominant pathway of *SLC6A1-*related neurodevelopmental disorders (NDDs), however, the molecular mechanism underlying the variable clinical presentation remains unclear.

**Methods:** Here, through work of the Undiagnosed Diseases Network, we identify an undiagnosed individual with an inherited p.(A334S) variant of uncertain significance. To resolve this case and better understand the variable expressivity with *SLC6A1*, we assess the phenotypes of the proband with a cohort of cases diagnosed with *SLC6A1-*related NDDs. We then create an allelic series in the *Drosophila melanogaster* to functionally characterize case variants.

**Results:** We identify significant clinical overlap between the unsolved case and confirmed cases of *SLC6A1-*related NDDs and find a mild to severe clinical presentation associated with missense variants. We confirm phenotypes in flies expressing *SLC6A1* variants consistent with a partial loss-of-function mechanism.

**Conclusion:** We conclude that the p.(A334S) variant is a hypomorphic allele and begin to elucidate the underlying variability in *SLC6A1*-related NDDs. These insights will inform clinical diagnosis, prognosis, treatment and inform therapeutic design for those living with *SLC6A1-* related NDDs.

## Introduction

*Solute Carrier 6, Member 1* (*SLC6A1*) encodes the γ-aminobutyric acid (GABA) transporter, type 1 (GAT-1) protein which is responsible for the reuptake of GABA from the synaptic cleft and for extracellular clearing^1^. *SLC6A1* has been linked to autosomal dominant myoclonic-atonic epilepsy (MAE, [MIM: 616421]) and was recently implicated in a novel disease, *SLC6A1*-related neurodevelopmental disorders. In 2020, clinical characterization of 116 cases involving *SLC6A1* identified epilepsy to be the most common symptom (91.1%) followed by developmental delay (82%)^2^. Additionally, variable clinical disease presentation of intellectual disability, movement disorder, language disorder, hypotonia, sleep disorders, autism spectrum disorder (ASD), and attention-deficit/hyperactivity disorder (ADHD), have been observed^2–5^. The wide phenotypic spectrum associated with *SLC6A1*-related disorders leads to prognostic uncertainty, and treatment is largely reactive, addressing symptoms as they arise. Furthermore, in many cases seizure management is unsuccessful indicating improved therapeutics are needed, however a thorough evaluation of the underlying molecular mechanisms contributing to *SLC6A1* pathology is required to enlist targeted therapeutics.

Notably, clinical reports have identified enrichment for *de novo* heterozygous missense variants as compared to truncating variants in *SLC6A1* among affected individuals^4,6^. The proposed molecular mechanism underlying *SLC6A1*-related neurodevelopmental disorders is haploinsufficiency, which is supported with a high probability of loss-of-function intolerance score (pLI=1, gnomAD v4). In 2024, GABA uptake was evaluated for 166 variants *in vitro* identifying 77 strong loss-of-function variants with severely impaired GABA clearance, 27 weak loss-of-function, and two gain-of-function variants with accelerated clearance^7^. This research thus confirmed that haploinsufficiency is the predominant mechanism responsible for *SLC6A1*-related neurodevelopmental disorders^7^. However, this study also concluded that 60 individuals affected with *SLC6A1*-related disorders have weak loss-of-function alleles that result in GABA reuptake in mildly reduced normal ranges. This suggests that GABA levels are tightly regulated, and GABA dysregulation has been linked to neurodevelopmental disorder and epilepsy pathogenesis^8^. GABA is the primary inhibitory neurotransmitter in the human cortex, and modulating GABA uptake and catabolism has long been a target of anti-epileptic drugs^9^. GATs have been used as therapeutic targets to modulate seizure phenotypes, and variants *SLC6A1* were recently been linked to human disease^10^.

Currently, over 100 cases of *SLC6A1*-related neurodevelopmental disorder have been identified^2^. Despite *SLC6A1* being well documented in disease pathology, variants of uncertain significance continue to pose a difficult diagnostic question. With established disease genes, thorough phenotypic and functional characterization of confirmed cases will improve the diagnostic journey of future affected individuals. Deep clinical phenotyping analysis of an undiagnosed case compared to a cohort of confirmed *SLC6A1* cases provides an opportunity to generate new hypotheses regarding involvement of *SLC6A1* variants in neurodevelopmental phenotypes. Leveraging *in vivo Drosophila* models not only allows confirmation of candidate genes and novel genetic variants, but also the evaluation of dysregulated neural development through phenotypic behavioral assessment. Furthermore, creation of an allelic series can be used to rank variant effect in a uniform genetic background. This is particularly important for variants that present with variable symptoms among affected individuals. In this study, we assess an undiagnosed individual harboring an inherited p.(A334S) variant of uncertain significance (VUS) through deep phenotypic and *in vivo* functional characterization.

## Materials and Methods

### Identification of individuals

Individual 1 (UF1) was referred to the UDN at Vanderbilt University by their local physician^11,12^. The case was referred to the model organisms screening center for evaluation as a variant of uncertain significance in a known disease associated gene (*SLC6A1* (NM_003042.4:c.1000G>T p.(A334S)). Additional individuals were identified through the National Brain Gene Registry (BGR, Washington University, St. Louis, MO, 63130 USA) and recruited to the study (BCM IRB of BCM gave ethical approval for this work : H-49443). *SLC6A1* variants were identified by epilepsy panel genetic testing, exome, or genome sequencing in a CAP/CLIA-certified diagnostic laboratory. The variant identified in UF1 was confirmed by Sanger sequencing.

### Clinical evaluation

Individual 1 (UF1) received a thorough clinical evaluation at the UDN Vanderbilt clinical site, which involved a multidisciplinary approach. The assessments included genetic testing and detailed phenotype evaluations conducted by multiple specialists^13^. To accurately evaluate the phenotypic variability associated with *SLC6A1* variants, we analyzed a cohort of 13 individuals with known pathogenic *SLC6A1* variants, all of whom were phenotyped using standardized method described in Baldridge, D. *et.al.,* 2024. Clinical data for these confirmed cases were obtained from the Brain Gene Registry Portal (https://braingeneregistry.wustl.edu/)^14^. All the phenotypic data for all the individuals in the study was collated and annotated with the Human Phenotype Ontology (HPO) terms using Doc2Hpo^15^ followed by manual curation. The number of HPO terms per proband assesses the depth of phenotyping in this cohort (Figure S1). The HPO terms were then categorized by disease concept domains to assess the most impacted domains.

Rapid neurodevelopmental assessment protocol (RNAP) evaluations were completed by clinicians, parents, or caregivers at National Brain Gene Registry (BGR, Washington University, St. Louis, MO, USA) recruitment sites. The 804-item questionnaire was categorized into seven domains including development, communication, seizure, motor, and sleep. In cases where a status of affected or unaffected by symptoms is reported, a value of 0=unaffected and 1=affected is assigned. Severity was assigned based on the number and severity of symptoms experienced by each individual on a 1-5 scale, where 1=mildly affected and 5=profoundly affected. A RNAP domain score was generated for each individual, which was then normalized based on the number of questions answered by the individual in each domain.

### Human Phenotype Ontology (HPO)-based quantitative phenotypic similarity analysis

A detailed description of the methods used for HPO-based quantitative phenotypic similarity analysis has been previously published^16^. Briefly, phenotype similarity scores are generated using Lin’s semantic similarity method, with the OntologyX suite of R packages. A minimum of four distinct HPO terms are required per individual for inclusion in analysis. The Lin similarity scores represent the similarities and differences in phenotypes (as represented by HPO terms), between two individuals. Pairwise similarity matrices are used to generate a distance matrix. Hierarchical agglomerative clustering (HAC) is performed on the distance matrix to identify various phenotypic clusters. A heatmap is generated with the ComplexHeatmap R package based on the previously generated similarity matrix and ordered according to HAC clustering.

### Fly stocks and husbandry

Fly stocks are housed at 25C and held in a 12-hour light/dark (L/D) cycle. Flies are removed from the incubator for the duration of time required for assessment and are not transferred to room temperature for more than 30 minutes.

Fly lines were obtained from the Bloomington *Drosophila* stock center (BDSC) and the Kyoto *Drosophila* stock center (DGRC).

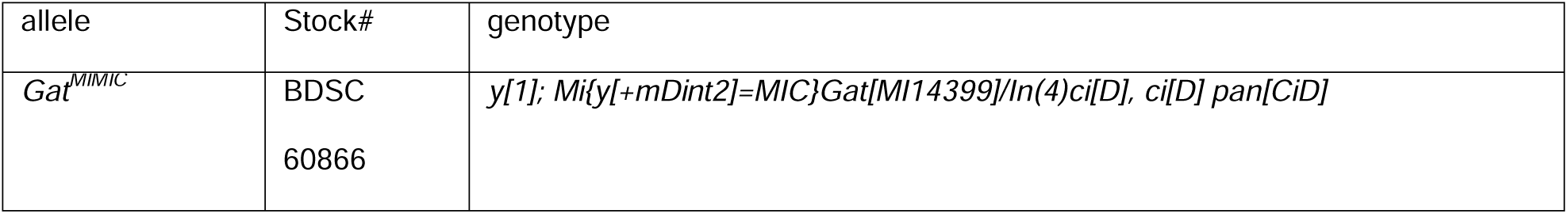
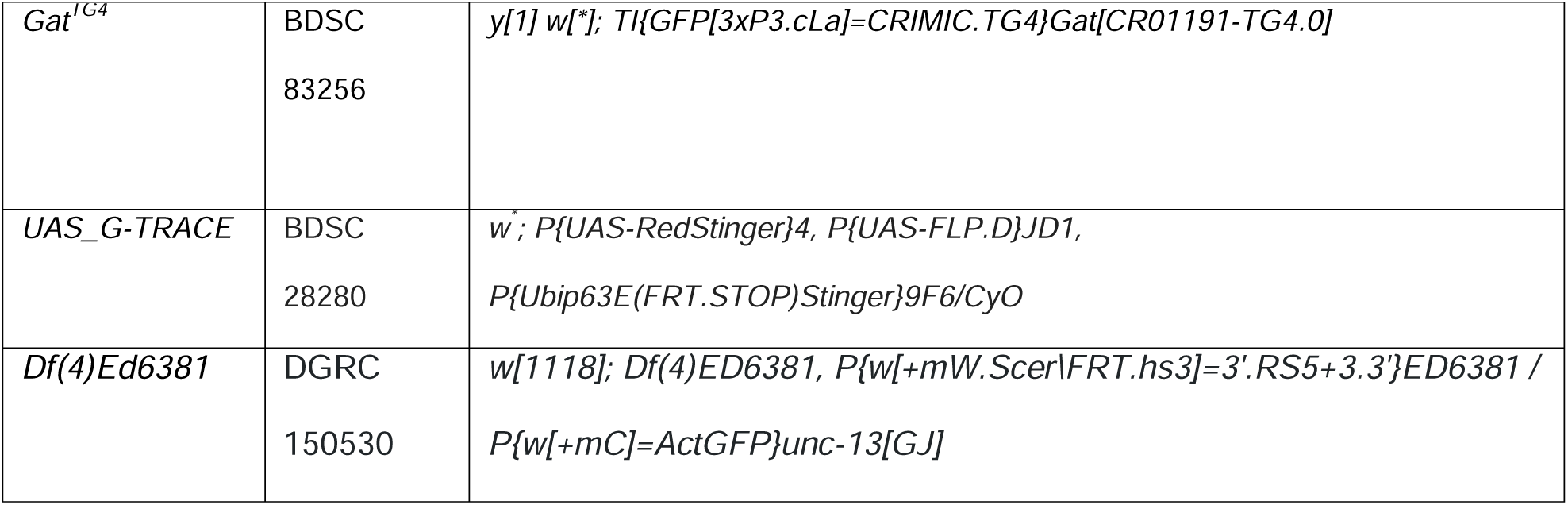

### Human *SLC6A1* transgenic generation

The human *SLC6A1* clone was obtained from the Kenneth Scott cDNA collection at Baylor College of Medicine. Variant lines are created using the New England Biolabs Q5 site directed mutagenesis kit for p.(A288V) (*SLC6A1^A288V^*), p.(S295L) (*SLC6A1^S295L^*), p.(G297R) (*SLC6A1^G297R^*), and p.(A334S) (*SLC6A1^A334S^*). Successful mutagenesis is confirmed by Sanger sequencing. Vectors are transformed into NEB 5-alpha competent cells (New England Biolabs, Ipswich, MA, USA). Gateway compatible donor vector constructs are converted to the pGW destination vector using LR clonase and clones are selected by antibiotic resistance. Plasmid containing the *SLC6A1* reference or variant cDNAs is confirmed by Sanger sequencing and injected into n>200 *PhiC31 ; VK0037* embryos at a concentration of 400 ng/µL. All constructs are injected into the same docking site (VK37) to avoid changes in expression level. Transgenic flies are crossed to the laboratory control strain *y^1^ w\** and integration of the cassette is selected for by expression of the mini white gene. Stocks are established by crossing to the SM6A balancer.

### *Drosophila* ortholog *dSLC6A1* transgenics generation

To create the *Drosophila Gat* transgenic lines, the wild-type *Gat* genomic sequence was extracted by PCR from the Canton S strain of *Drosophila.* GoldenBraid 2.0 cloning was completed as previously described^17,18^. Briefly, *Gat* genomic fragments and attB sites are synthesized and inserted into GoldenBraid 2.0 compatible pUPD2 constructs. pUPD2 fragments are converted to the alpha-1 destination vector and transformed into maximum efficiency DH10B competent cells (Thermo Fisher Scientific, Houston, TX, USA). To create the *SLC6A1* p.(A334S) homologous variant p.(A372S) in the fly ortholog *Gat*, mutagenesis primers are designed using NEBaseChanger (nebasechanger.com) and PCR mutagenesis is completed with the Q5 site directed mutagenesis kit (New England Biolabs, Ipswich, MA, USA). Vectors are transformed into NEB 5-alpha competent cells (New England Biolabs, Ipswich, MA, USA). Confirmation of the variant and genomic sequence is completed by Sanger sequencing. Constructs are co-injected at a concentration of 400 ng/µL with PhiC-31 (gift from Dr. Hugo Bellen) into *Gat^TG4^* (n>200) embryos to insert the constructs into the *Gat* locus. Transgenic flies are crossed to the fourth chromosome balancer *TI{TI}Crk[dsRed]^12^/In(4)ci[D], ci[D] pan[ciD]* (BDSC:90851) and incorporation of the construct is confirmed by loss of the EGFP marker on *Gat^TG4^*. To create the *Gat* transgenic wild-type (thereafter, transgenic *Drosophila SLC6A1 dSLC6A1*) and p.(A372S) (thereafter, *dSLC6A1^A334S^*) stocks, n>2 lines are established by sibling cross.

### Lineage tracing and brain immunostaining

Current *Gat^TG4^* expression is determined by crossing to the *UAS-GTRACE* line and screening for RFP positive progeny. Larval brain dissection is performed, and larval brain preps are fixed in ice cold 4% PFA/PBS/2% Triton overnight. Brains are blocked in PBS/2%Triton/5% normal donkey serum for 1 hour. Primary antibodies are incubated overnight; mouse anti-repo (Developmental Studies Hybridoma Bank, IA, USA Cat# 8D12, 1:50). Secondary antibodies are incubated for two-hours at room temperature; mouse anti-Cy3 (1:250). Brains are mounted and imaged using a Zeiss 710 confocal microscope (Neurovisualization core, Baylor College of Medicine, TX, USA) with 1 μm sections.

### Seizure sensitivity evaluation

Flies are isolated in a single culture vial without food and are subjected to a ten second mechanical vortex (Fisher STD Vortex Mixer, Cat. No, 02215365). Duration of initial seizure, a variable period of paralysis, and recovery seizure is recorded for n>20 adult flies at 10-12 days after eclosion.

### Lifespan assay

Groups of 10 flies of each genotype are placed together in a culture tube, collecting a total of 100 flies per genotype. Flies are collected daily to confirm date of eclosion. Survival is evaluated every 3-4 days and food changed to minimize bacterial accumulation confounding the assay. Duration of overall lifespan is evaluated, and Kaplan Meier curves and a Log-rank Mantel Cox test is used to determine significant differences between groups.

### Sleep and activity assessment

Seven-to-ten-day-old flies (n>15) are loaded into the *Drosophila* Activity Monitor (DAM, Trikinetics) and held under the same L/D cycle as their previous husbandry. Activity and sleep data per minute are recorded for seven days and periods of inactivity greater than five minutes are determined to be sleep periods. Sleep latency is the duration of time required to initiate sleep after lights out in the evening. Sleep onset is determined by calculating the duration in time to initiate sleep after lights out. After seven days in the DAM data are evaluated with automated strategies using MATLAB SleepMat software^19^. In human cDNA studies, variant lines are compared to expression of variant *SLC6A1* cDNAs. For variant knock-in studies, missense variant expressing lines are compared to a transgenic insert of the wild-type *Gat* genomic sequence.

### Statistical analysis

Statistical analysis is completed using GraphPad prism (Version 9.0.0). Continuous analysis is completed by Ordinary one-way ANOVA with Welch’s correction where differences between groups are quantified and a p-value less than 0.05 is considered significant.

## Results

### UDN proband clinical presentation is similar to individuals with confirmed *SLC6A1*-related neurodevelopmental disorder

Individual 1 (UF1) was referred for epilepsy, intellectual disability, and ASD. She faced delayed language development which further regressed along with loss of eye contact. Her seizures are not successfully managed by her current therapeutics. Family history is notable for neurodevelopmental disorders. Diagnostic assessments revealed normal chromosomal microarray, fragile X repeat analysis, plasma amino acid levels, acylcarnitine profile, and urine organic acids. Electroencephalogram recordings were consistent with Lennox Gastaut syndrome. Genetic testing of a 187-epilepsy gene panel (Invitae, CA, USA) revealed a variant in *SLC6A1* (NM_003042.4:c.1000G>T p.(A334S)). In gnomAD v4.10, this variant is absent, and the missense Z-score is 4.93 for the gene. Another variant at this amino acid position, NM_003042.4:c.1000G>C p.(A334P) has been reported as pathogenic (ClinVar ID: 192370). The present p.(A334S) variant was classified as a variant of uncertain significance (VUS) based on meeting PM5, PM2_supporting and PP2 criteria^20^. Family variant testing confirmed the p.(A334S) variant was maternally inherited. Exome sequencing and Intellectual Disability, Epilepsy & Autism (IDEA) panel testing identified the variant to be shared with her siblings that were similarly affected (**Figure 1A**). A diagnosis of catatonia recently was given to the proband and her younger brother. In summary, the family presented with an inherited variant in the *SLC6A1* gene, a locus more typically associated with *de novo* events leading to severe epilepsy. In order to determine if this variant was causative for the more variable presentation in the family, we pursued clinical and functional studies of the variant.

**Figure 1.**
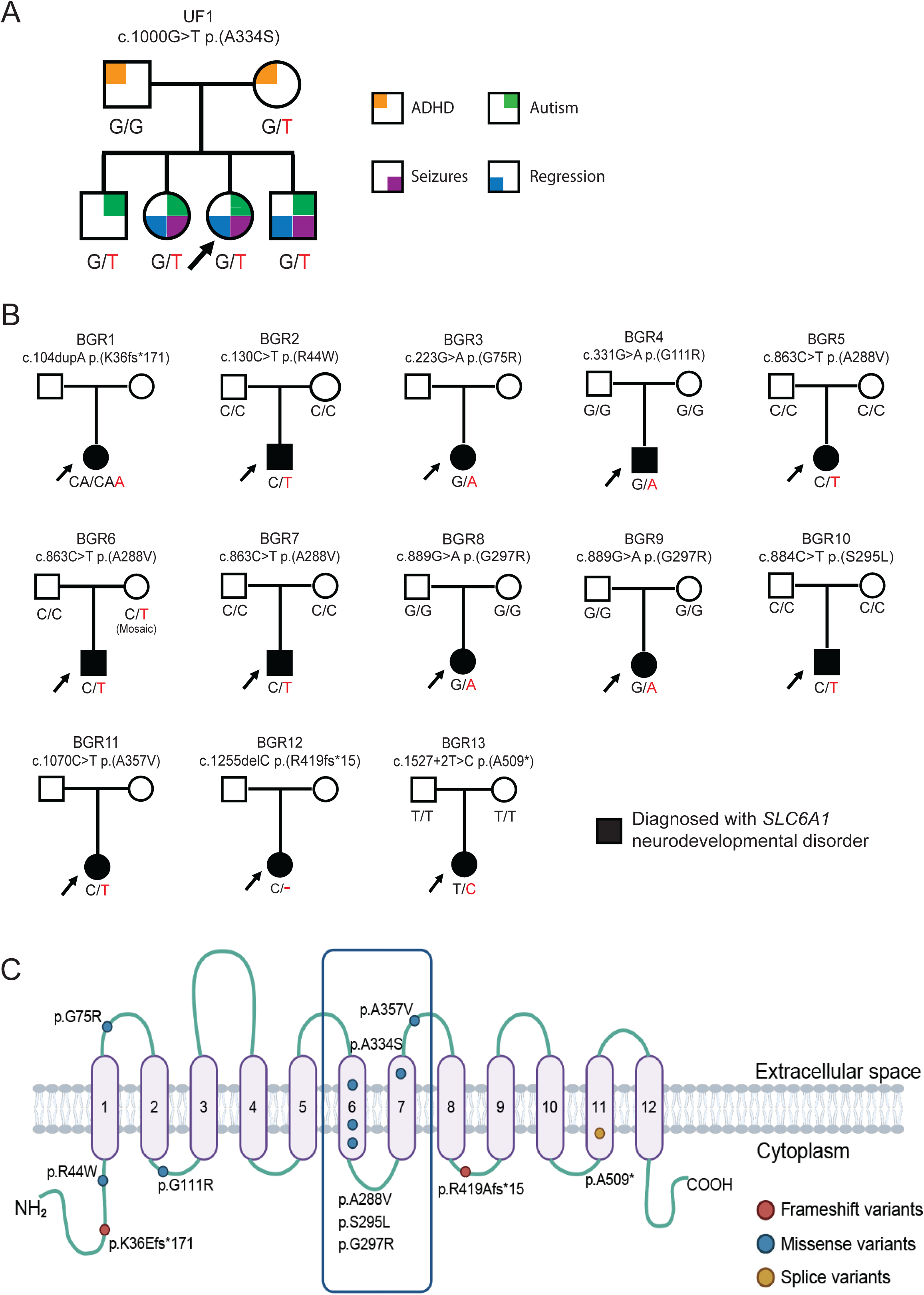
Comprehensive pedigrees and their genotypes for families with *SLC6A1* variants. Standard pedigree structures are utilized—filled circles and squares denote clinically affected individuals, and probands are indicated by black arrows. (A) The UF1 pedigree is denoted with the phenotypes as noted in family members respectively. (B) Thirteen families diagnosed with *SLC6A1-*neurodevelopmental disorders recruited through the Brain Gene Registry. (C) Schematic diagram of SLC6A1 protein structure from conceptual translation of transcript NM_003042.3 with mapping location (circles) of the amino acid variants observed in this study. In total, we identified eleven different SNVs and indels that are distributed throughout the protein. Blue circles represent missense variants, red circle represent frameshift variants and yellow circle represents the splice variant observed in this study. Image created in Biorender.

### Deep clinical phenotyping identifies the UDN candidate p.(A334S) variant is associated with neurological, cognitive, and communication impairment

To better assess whether the UDN candidate p.(A334S) VUS variant is linked to a more variable presentation within the family, we examined a cohort of 13 individuals with known pathogenic variants in *SLC6A1*. We recruited thirteen individuals with known *SLC6A1* variants to the Brain Gene Registry, a research study aimed to provide detailed phenotypic data for individuals with variants in known neurodevelopmental genes. The clinical data were accessed through the BGR portal (https://braingeneregistry.wustl.edu/). Reported *SLC6A1* variants are heterozygous in each of these known *SLC6A1* individuals, and a majority (8/9 for which both parental samples were available) arose *de novo* (**Figure 1B**). When we map the confirmed BGR variants p.(K36Efs*171), p.(R44W), p.(G75R), p.(A288V), p.(S295L), p.(G297R), p.(A357V), p.(R419Afs*15), p.(A509*) and candidate UDN variant p.(A334S) to the GAT-1 protein, there is an enrichment for variants in the 6^th^ and 7^th^ transmembrane domains, which are part of the active pore to facilitate GABA transport (**Figure 1C**, Table S1). Importantly, the p.A334S variant lies in transmembrane domain 7, and the p.(A334P) variant was determined to be a strong loss-of-function allele *in vivo*^7^. Deep clinical phenotyping of these individuals (on average 12 HPO terms per individual) demonstrates the wide spectrum of severity of clinical features observed (Figure S1), making this cohort an ideal dataset for comparison to the UF1 proband and family members. Recently Goodspeed *et.al.* proposed the draft conceptual disease model that identifies key domains of disease describing the phenotypes of affected individuals with *SLC6A1* neurodevelopmental disorder^21^. Based on the HPO terms annotated for each individual in our cohort, we classified them under 13 domains including neurological, motor, cognitive, developmental, communication, behavioral, emotional, gastrointestinal, visual, sleep, musculoskeletal, growth and other. Neurological and cognitive domains are severely impacted in our cohort. Seizures are managed with general anti-epileptics with varying levels of success; however, cognitive impairment persists regardless of seizure suppression. Additionally, we note domains based on clinical data such as high incidence of behavioral, motor, developmental, and communication symptoms which could further exacerbate the disease’s burden on daily life for individuals (**Figure 2A**). Increased support in these areas could improve *SLC6A1* symptoms for affected individuals in the near term, and GAT-1 targeted therapeutics could potentially alleviate these symptoms in the extended future.

**Figure 2.**
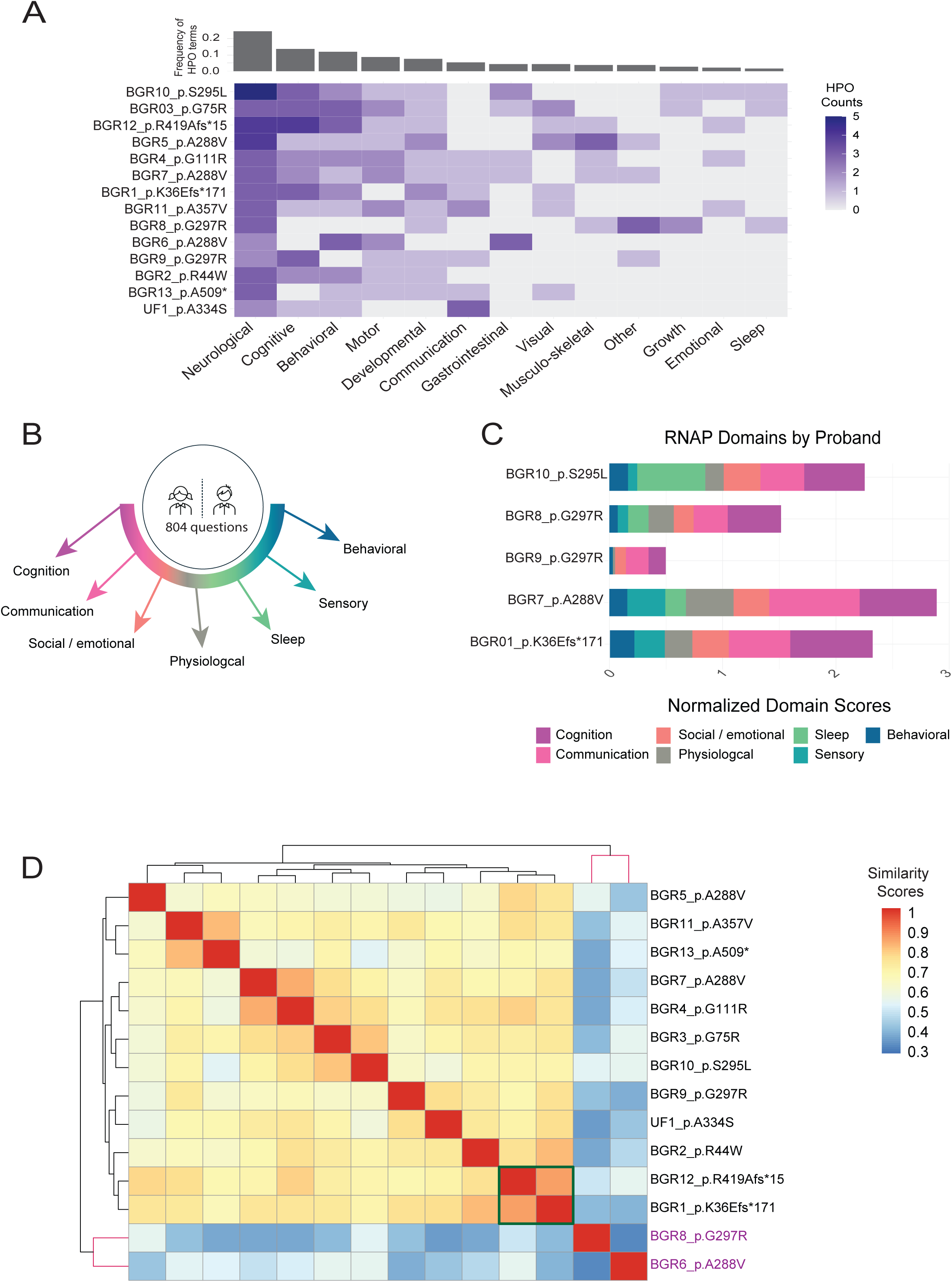
Clinical phenotypic analyses. (A) *SLC6A1* phenotypic grid of all individuals in the study. HPO terms from all individuals were categorized into 11 key phenotypic domains described in the draft *SLC6A1* disease model. The frequency of terms per phenotypic domain is plotted. Neurological and cognitive domains show the highest frequency of terms. (B, C) Rapid neurodevelopmental assessment protocol (RNAP) profiles of individuals. The 804-item questionnaire in the RNAP was categorized into 7 domains namely: cognitive ability, adaptive behavioral function, social and emotional function, sensory-motor function, sleep symptoms, communication ability, physical features and neurologic symptoms. The RNAP domain score was generated for each individual, which was then normalized based on the number of questions answered by the individual in each domain. These scores are plotted to visualize the RNAP profile for each individual. (D) Phenotypic similarity heatmap for individuals with *SLC6A1* variants. A heatmap was generated using proband phenotypic similarity scores and ordered based on Hierarchical Agglomerative Clustering of proband phenotypic similarity. Dendrograms showing clusters are present at the left and top sides of the heatmap. Green box highlights the two frameshift variants that cluster together with similar severe phenotypes. The UDN p.(A334S) variant clusters with other confirmed cases of *SLC6A1*.

As part of the BGR assessment, we evaluated the neurodevelopment of individuals with *SLC6A1* variants using the rapid neurodevelopmental assessment protocol (RNAP)^14^. This standardized neurocognitive assessment includes domains including cognitive ability, adaptive behavioral function, social and emotional function, sensory-motor function, sleep symptoms, communication, physical features, and neurologic symptoms. The 804-item questionnaire survey was evaluated in these seven domains (**Figure 2B**). A RNAP domain score was generated for each individual (Figure S2). Normalizing the scores with the number of questions answered by the individual in each domain, we can visualize a phenotypic profile for each individual (**Figure 2C**). Cognition and communication abilities are the two domains that are most affected. These critical domains are also identified using HPO analysis based on available clinical data, further emphasizing these as dominant symptoms of *SLC6A1.* Overall, this clinical assessment from the BGR gave a reasonable dataset for understanding the impact of the full spectrum and the severity of *SLC6A1*-neurodevelopmental disorders, and therefore an excellent comparison set for studying the variant of uncertain significance from the UF1.

### *SLC6A1* UDN p.(A334S) variant clusters phenotypically among confirmed *SLC6A1*-related neurodevelopmental disorder cases

Through HPO-based phenotypic clustering analysis we can systematically compare and interpret the clinical manifestations of the UF1 and the BGR cases, facilitating the assessment of the variant disease relevance and guiding further investigation and diagnosis. Phenotypic similarity scores generated using the Lin similarity method are calculated and visualized in a heatmap (**Figure 2D**). Importantly, we find that the symptomology of the UF1 bears a significant similarity to other *SLC6A1*-affected individuals. UF1 clusters closely with BGR9_p.G297R (**Figure 2D**), and these cases have a similar phenotypic profile including neurological, cognitive, and communication deficits (**Figure 2A**). Additionally, the recurrent p.(G297R) variant is also subject to variable expressivity suggesting a mild to severe variable presentation shared by these variants.

This analysis was not only informative for the UDN case in question, we also note, even with highly similar phenotypic scores associated with *SLC6A1* variant alleles, subclusters of related phenotypic profiles can be identified. The two individuals with frameshift variants in the BGR cohort cluster together with consistent severe phenotypes including global developmental delay, EEG abnormalities, tremors, and language impairment. Individuals harboring *SLC6A1* variants that have been shown to have strong loss of function variants (p.(S295L), p.(A288V), p.(G111R), p.(G75R)) cluster together with phenotypes such as autistic behavior, EEG abnormalities, aggressive/impulsive or violent behavior, and memory impairment. Through this analysis, we also observe phenotypic variability among recurrent missense variants (p.(A288V) and p.(G297R)) that suggests additional factors may be influencing symptom presentation. For example, the individual listed as BGR8_p.G297R does not cluster with individual BGR9_p.G297R even though they share the same variant. Dissection of BGR8_p.G297R proband’s phenotypes suggests it to be a case of dual molecular diagnosis, with a combination of the traits observed resulting from both the *SLC6A1* variant and the *PTPN11* variant (Figure S3). Moreover, BGR6_p.A288V, who harbors a strong loss-of-function variant, has additional severe gastrointestinal phenotypes that skew the clustering analysis. It is important to note that this individual has a history of localized-idiopathic seizures, paroxysmal dyskinesia, extra-axial cerebrospinal fluid accumulation, tremor, chorea, abnormality of movement, hyperactivity, impulsivity, and gastroesophageal reflux. It is challenging to discern whether the gastrointestinal symptoms are caused by *SLC6A1* given the unique symptomology in this case. Comprehensive documentation of gastrointestinal phenotypes during individual evaluation will clarify whether gastrointestinal phenotypes are a previously unrecognized component of *SLC6A1*-neurodevelopmental disorders. The deep phenotypic characterization along with the quantitative phenotypic clustering analysis allows us to map the severity and breadth of clinical features across individuals and to elucidate gene- and allele-specific phenotypic differences. We were expecting to uncover a genotype-phenotype correlation; however, the variability in phenotypes observed especially with the missense variants, warrants functional characterization.

### Expression of *SLC6A1* suppresses mild seizure phenotypes in *Drosophila*

Next, we sought to create an allelic series to rank the strength of each allele *in vivo.* To compare the function of the UDN p.(A334S) variant to the confirmed BGR variants in a uniform genetic background, we created variant specific constructs and expressed them in *Drosophila.* We utilized the GAL-4/UAS binary expression system to express human cDNAs in critical tissues in flies. To avoid off-target effects we drove expression of our *SLC6A1* alleles under the promoter of the homologous fly gene. *SLC6A1* is orthologous to *Gat* in flies with high sequence identity (342/621-55%) and similarity scores (446/621-71%)^22^*. Gat* is known to be expressed in fly glia to facilitate GABA uptake in a similar recycling pathway to humans GABA recycling^23^. Previous research identified that *Gat* mutants are capable of suppressing seizure-like activity in flies, post-kinetic shock consistent with increased GABA-ergic signaling^24,25^. We obtained the *Gat* specific Trojan GAL-4 fly line (*Gat^TG4^*) which is a strong loss-of-function allele^26^. The *Gat^TG4^* cassette is inserted into the first coding intron of *Gat* and contains a splice donor, T2A ribosomal skipping sequence, GAL-4 transcriptional activator, and a *poly-A* signal (**Figure 3A**). This results in severe premature truncation of the *Drosophila* protein and simultaneously expresses a GAL-4 in the same spatial and temporal expression pattern as the *Gat* gene. This allele can be used to drive expression of an upstream activation sequence (*UAS*) containing construct to identify expression pattern or to express the human cDNA in the fly. *Gat* is homologous to the GAT encoding *SLC6A1/11/12/13* genes in humans, so variant expression could result in more pronounced phenotypes in the fly. We created transgenic lines harboring the human reference (*SLC6A1^Ref^*), UDN variant p.(A334S) (*SLC6A1^A334S^*), strong loss of function BGR variants p.(A288V) (*SLC6A1^A288V^*), p.(S295L) (*SLC6A1^S295L^*), and recurrent variant p.(G297R) (*SLC6A1^G297R^*) constructs preceded by a *UAS* sequence inserted into the same genomic locus. Critically, all variants are conserved between humans and flies and therefore potentially serve a conserved function (**Figure 3B**). In humans, *SLC6A1* is expressed in both neurons and glia, however astrocytic dysregulation of GAT-1 is believed to underlie *SLC6A1* pathology^27^. In flies, *Gat* is expressed exclusively in glia^23^. First, we confirmed that our *Gat^TG4^* allele is expressed only in repo positive glial cells to ensure proper expression of the *SLC6A1* reference and variant lines (**Figure 3C-E**).

**Figure 3.**
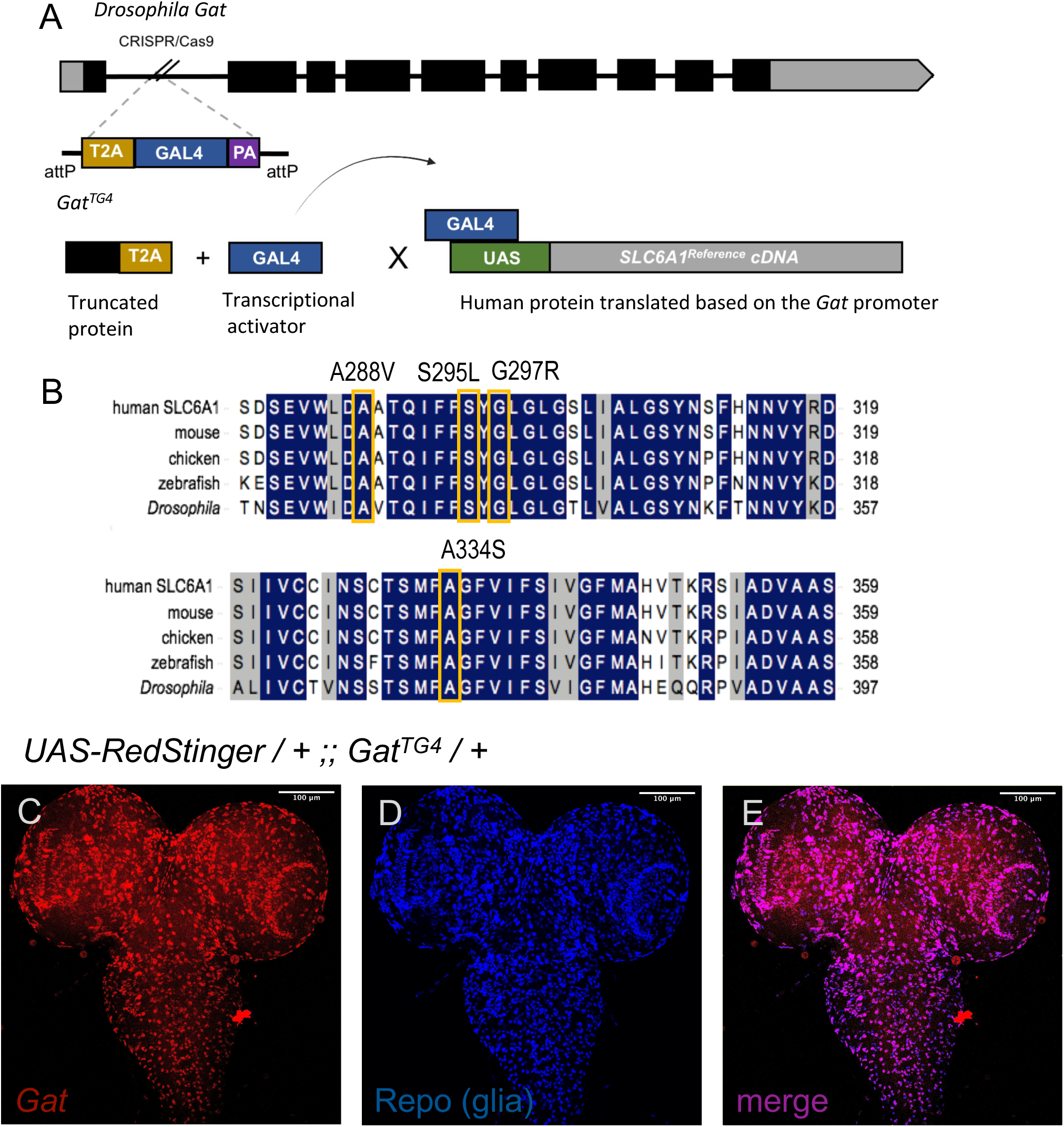
*Gat* specific *GAL-4* transcriptional activator enables spatial and temporal control of *SLC6A1* variant expression. (A) The Trojan GAL-4 cassette is a strong loss-of-function allele that simultaneously truncates the *Drosophila* Gat protein and expresses a GAL-4 protein in under the regulatory control of the fly gene promoter. (B) Upstream activation sequence containing constructs were generated with the UDN and BGR variants p.(A288V), p.(S295L), p.(G297R), and p.(A334S) for functional analysis in the fly model. (C) Current lineage tracing identifies *Gat^TG4^* expression in the fly larval brain. (D) anti-Repo immunostaining labels glial cells. (E) *Gat* and Repo merge identifies that *Gat^TG4^* is expressed in Repo-positive glial cells.

Next, we evaluated seizure phenotypes in adult flies. Seizures are a prominent feature of *SLC6A1* disease and previous *SLC6A1* studies identified temperature sensitive seizures in variant expressing flies^28^. We enlisted the bang sensitivity assay to test induced seizure recovery^29^. Wild-type flies do not experience mechanical stimulus induced seizures, however, fly mutants that are sensitive to seizures exhibit delayed seizure recovery. There is immediate recovery and no seizure sensitivity induced by mechanical vortexing in the background control strain *y^1^ w*.* There is also no bang sensitivity observed in 10-day-old flies on a wild-type background when *SLC6A1^Ref^* or any variant line which includes both the known *SLC6A1* variants from the BGR cases and the VUS from the UF1 is expressed (Figure S4A). We then tested overexpression of *SLC6A1^Ref^* in a *Gat* null background to evaluate the ability to the human reference cDNA to rescue loss of the fly gene. *Gat* null animals were created by pairing *Gat^TG4^* with a deficiency line that spans the *Gat* locus (*Df(4)Ed6381*). We identified that loss of *Gat* results in bang sensitive animals. We performed a human cDNA rescue paradigm by driving expression of the human reference cDNA in a fly null genetic background (*Gat^TG4^ / Df(4)Ed6381*). Expression of *SLC6A1^Ref^*fully rescues bang sensitivity (Figure S4B). Bang sensitive mutants have also been associated with reduced overall longevity so we evaluated overall lifespan when *SLC6A1^Ref^* is expressed^30^. Loss of *Gat* also results in reduced longevity in a dosage sensitive manner when *Gat^TG4^* is expressed in a heterozygous and homozygous state (Figure S4C). Interestingly, expression of *SLC6A1^Ref^* with *Gat^TG4^* results in more severe reduced longevity than *Gat* null animals (Figure S4C). Since this is occurring in a wild-type background using induced overexpression, these results indicate that *Gat* could be a dosage sensitive gene, and therefore both gain- and loss-of-function variants could be detrimental.

### UDN variant p.(A334S) induces loss-of-function phenotypes including severely reduced sleep latency

Since we observed no bang sensitivity, we sought another method to evaluate our variants behaviorally. The role of GABA in the regulation of sleep is well established. Circadian rhythm regulates sleep and activity behavior to maintain sleep and wake cycles in mammals and invertebrates^31^. In flies, GABA receptor recruitment to the plasma membrane is increased to inhibit excitatory clock neurons to promote sleep. The role of *Gat* in this regulatory loop was recently established using a hypomorphic allele of *Gat^25^*. Phenotypes associated with *Gat* loss-of-function include severe reduced sleep latency, increased overall sleep, decreased sleep bout number, and increased sleep bout duration^25^. *Gat* mutants transition to sleep rapidly at dusk (or lights out in a laboratory environment) and when they initiate a sleep bout, they remain asleep for an extended duration and thus experience fewer overall sleep bouts. Changes in sleep latency and total sleep have also been observed in variant forms of the GABA_A_ receptor *Resistant to dieldrin* (*Rdl*) in flies further supporting the role of GABA in sleep regulation^32^.

We found that heterozygous *Gat^TG4^* animals exhibit a drastic decrease in sleep latency. This is suppressed by expression of the human reference *SLC6A1^Ref^*to wild-type levels as seen with the laboratory control strain *y^1^ w*.* Human *SLC6A1* variant lines (both known and candidate UDN VUS) exhibit a severe reduction in sleep latency similar to *Gat^TG4^*. Animals expressing *SLC6A1^A288V^*, *SLC6A1^S295L^*, *SLC6A1^G297R^*, or *SLC6A1^A334S^*, all experience reduced sleep latency compared to *SLC6A1^Ref^* but are less significantly impacted than *Gat^TG4^ / +* which is expected with partial loss-of-function variants (**Figure 4A**). There is a decrease in overall sleep when *SLC6A1^G297R^*or *SLC6A1^A334S^*is expressed compared to *SLC6A1^Ref^* (**Figure 4B**). Interestingly, when *SLC6A1^Ref^* is expressed there is an overall increased sleep consolidation and for all variants sleep is more fragmented with a decrease in sleep bout length and an increase in sleep bout number (**Figure 4C-D**). All variants fail to induce increased sleep consolidation in both sleep bout length (**Figure 4C**) and bout number (**Figure 4D**). This could be due to decreased sleep latency allowing flies to experience a greater number of sleep bouts and therefore each is of shorter duration. There is also an increase in overall daily activity when *SLC6A1^S295L^*, *SLC6A1^G297R^*, or *SLC6A1^A334S^*is expressed (**Figure 4E**). Glial expression of human *SLC6A1* suppresses reduced sleep latency in flies but also induces abnormal phenotypes in additional facets of sleep behavior. Expression of any *SLC6A1* variants, including the UDN candidate variant p.(A334S), fail to recapitulate *SLC6A1^Ref^* induced sleep phenotypes indicative of a hypomorphic loss-of-function mechanism.

**Figure 4.**
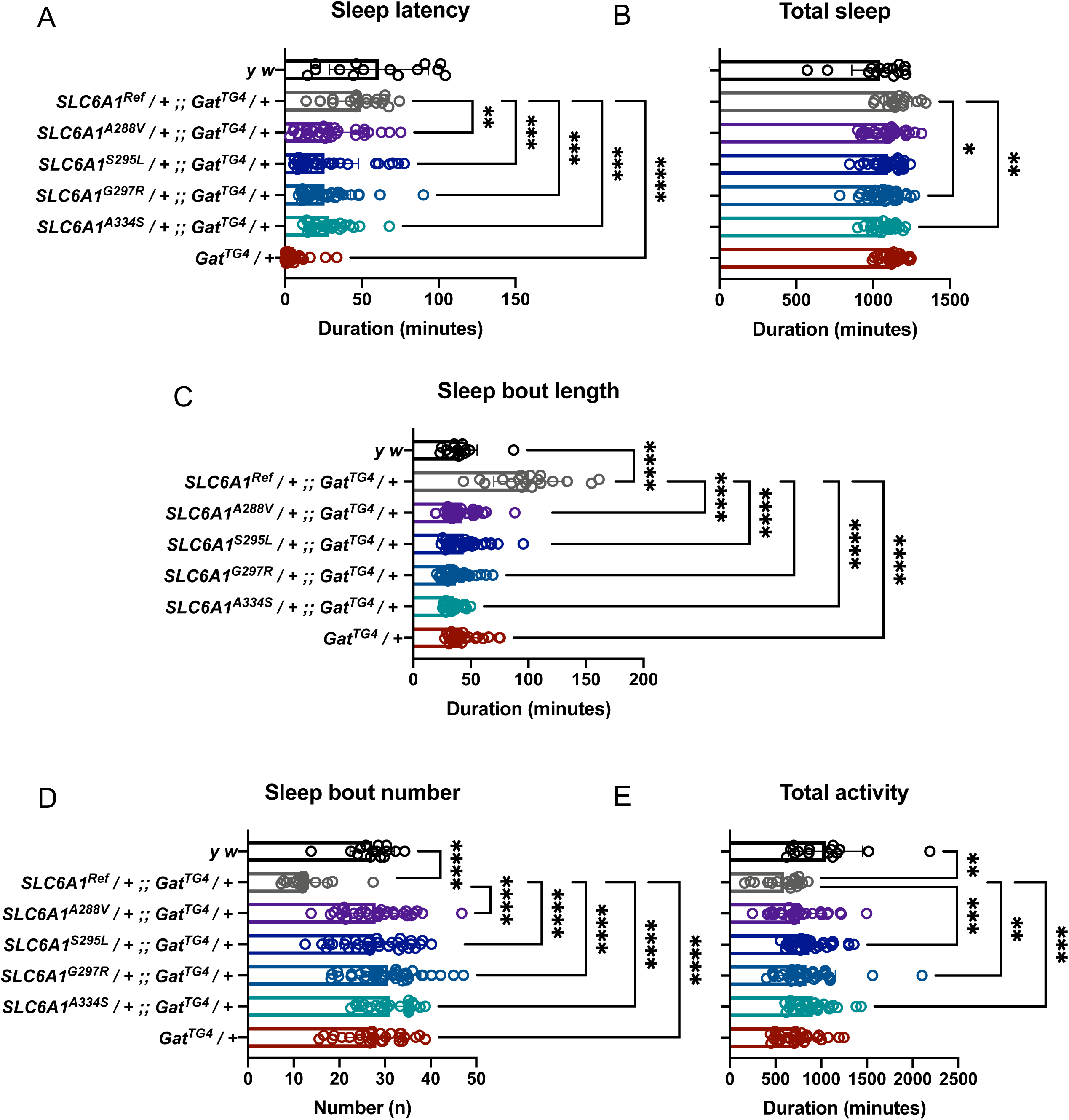
Expression of *SLC6A1^A334S^* results in decreased sleep latency. (A) Expression of a heterozygous loss-of-function allele *Gat^TG4^ / +* results in a severe decrease in sleep latency compared to *SLC6A1^Ref^ ;; Gat^TG4^ / +* (P<0.0001). Sleep latency is suppressed by expression of *SLC6A1^Ref^* driven by *Gat^TG4^*. Expression of variant lines also induces a significant decrease in sleep latency *SLC6A1^A288V^* (P=0.067), *SLC6A1^S295L^* (P=0.0006), *SLC6A1^G297R^* (P=0.0001), *SLC6A1^A334S^* (P=0.0008) compared to *SLC6A1^Ref^.* (B) Total sleep is reduced for *SLC6A1^G297R^* (P=0.0141) and *SLC6A1^A334S^* (P=0.0064). (C) Sleep bout length is reduced in all variants *SLC6A1^A288V^* (P<0.0001), *SLC6A1^S295L^* (P<0.0001), *SLC6A1^G297R^* (P<0.0001), *SLC6A1^A334S^* (P<0.0001). (D) Sleep bout number is increased for all variants *SLC6A1^A288V^* (P<0.0001), *SLC6A1^S295L^* (P<0.0001), *SLC6A1^G297R^* (P<0.0001), *SLC6A1^A334S^* (P<0.0001). (E) Total activity is increased in *SLC6A1^S295L^* (P=0.0004), *SLC6A1^G297R^* (P=0.00075), *SLC6A1^A334S^* (P=0.0002). Welch’s ANOVA with Dunnett’s T3 multiple comparisons test.

We can further dissect these sleep and activity disruptions by evaluating behavior during the day and night. Overall sleep during the day is the only aspect analyzed where there is no phenotype induced by expression of the reference cDNA or any of the variants, but *SLC6A1^A288V^*, *SLC6A1^G297R^*, *SLC6A1^A334S^* all exhibit a decrease in total night sleep (Figure S5A). Fragmented sleep is consistently displayed by a majority of the variants including *SLC6A1^A334S^*with an increase in bout number and decrease in bout length during both the day and night (Figure S5B-C) and night activity is more significantly affected than day activity (Figure S5D). Of note, these results indicate that *Gat* loss-of-function induces reduced sleep latency and that expression of the human reference *SLC6A1* can suppress this. Expression of the UDN variant p.(A334S) results in a significant reduction in sleep latency indicative of a hypomorphic loss-of-function mechanism. Similar sleep phenotypes are also seen from the UDN and BGR variants p.(A288V), p.(S295L), p.(G297R), and these data are consistent with similar phenotypic clustering of the clinical data. Importantly, sleep latency identifies p.(S295L), p.(G297R), and p.(A334S) as stronger alleles and p.(A288V) as a weaker loss-of-function allele in flies.

### Expression of the p.(A334S) homologous variant in the *Drosophila SLC6A1* ortholog (*dSLC6A1*) also induces loss-of-function phenotypes

Using the overexpression of the human gene allowed us to build evidence for an impact on sleep for both the known *SLC6A1* pathogenic variants and the VUS from the UDN case. However, we did observe some evidence for toxicity from overexpression of the human gene. We therefore sought to further study the UDN variant p.(A334S) which is conserved between humans and flies (**Figure 3B**) in the context of the fly gene. To do this, we exchanged the swappable insertion cassette *Gat^TG4^* with either the wild-type (*dSLC6A1*) or p.(A334S) (*dSLC6A1^A334S^*) variant form of the orthologous *Drosophila* gene sequence (**Figure 5A**). This results in heterozygous variant expression as occurs in affected individuals. We used a heterozygous *Gat* loss-of-function control (*Mi{MIC}Gat[MI14399],* thereafter *Gat^MIMIC^*) that is lethal when crossed a deficiency line (*Df(4)Ed6381*). When we perform the bang sensitivity assay, we observe bang sensitivity from both the loss-of-function control *Gat^MImic^* and *dSLC6A1^A334S^*(**Figure 5B**), supporting a loss-of-function mechanism for p.(A334S).

**Figure 5.**
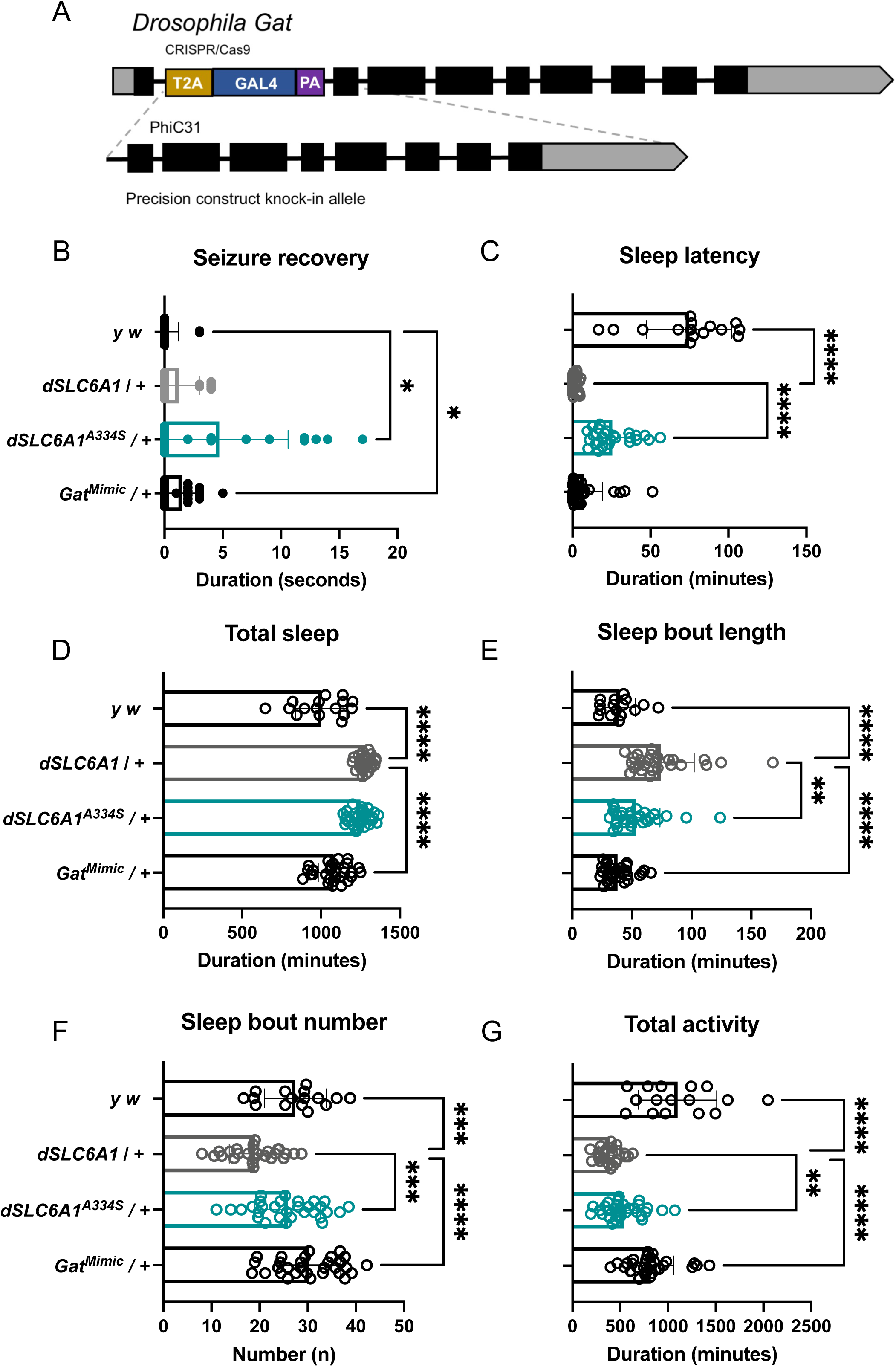
The UDN variant induces hypomorphic sleep phenotypes compared to expression of *dSLC6A1*. (A) Approach to replace the *Gat^TG4^* swappable insertion cassette with the *Drosophila Gat* sequence. This results in heterozygous expression of the transgene. (B) Expression of the loss-of-function control *Gat^MIMIC^* (P=0.0121) and *dSLC6A1^A334S^*(P=0.0110) results in seizure sensitivity compared to *y^1^ w\**. (C) There is a severe reduction in sleep latency when *dSLC6A1* is expressed and there is a significant increase in sleep latency in *dSLC6A1^A334S^* (P<0.0001) compared to *dSLC6A1.* (D) There is no significant difference between *dSLC6A1* and *dSLC6A1^A334S^*in total sleep. (E) There is a decrease in *dSLC6A1^A334S^* sleep bout length (P=0.0085). (F) There is an increase in *dSLC6A1^A334S^* sleep bout number (P=0.0003). (A) There is an increase in *dSLC6A1^A334S^* total activity (P=0.0043). Welch’s ANOVA with Dunnett’s T3 multiple comparisons test.

We note that overexpression of the human gene did not produce bang sensitivity. It seems likely that we observe this phenotype more sensitively in the context of the fly gene. Because the sleep assays were a more sensitive assay to capture hypomorphic alleles for the human *SLC6A1* constructs, we performed sleep and activity assessments using the fly construct lines. When we evaluate sleep, we observe severe decreased sleep latency when the loss-of-function control *Gat^MIMIC^*, or experimental *dSLC6A1* is expressed in a heterozygous fashion compared to *y^1^ w\** controls. There is however a significant increase in sleep latency when *dSLC6A1^A334S^*is expressed compared to *dSLC6A1* (**Figure 5C**). Similar to what is seen when the human *SLC6A1* is expressed, there is an overall increase in total sleep (**Figure 5D**), an increased sleep bout length (**Figure 5E**), and decreased bout number (**Figure 5F**) when *dSLC6A1* is expressed. Expression of *dSLC6A1^A334S^*also induces an increase in total sleep (**Figure 5D**), but *dSLC6A1^A334S^* fails to promote sleep consolidation to the extent of *dSLC6A1* and performs as an intermediate hypomorphic allele in sleep bout length and bout number (**Figure 5E-F**). Total activity is also impacted, with a significant decrease when *dSLC6A1* or *dSLC6A1^A334S^*is expressed (**Figure 5G**).

Again, expression of the wild-type transgenic control *dSLC6A1* induces phenotypes in every facet of sleep and activity during both the day and night (Figure S6A-D). However, whereas expression of the human *SLC6A1* variants induced more fragmented sleep, expression of the homologous p.(A334S) variant promotes sleep consolidation during the night similar to what is expected from loss-of-function variants. The *dSLC6A1^A334S^* variant induces a decrease in total day sleep and an increase in total night sleep (Figure S6A). The p.(A334S) variant also promotes night sleep consolidation with an increase in sleep bouts during the day of a shorter length, and no change in the number of sleep bouts during the night with a longer bout length compared to *dSLC6A1* (Figure S6B-C). There is also an increase in total day activity and a decrease on total night activity consistent with increased sleep during the night (Figure S6D). In this assay we see significant differences in behavior when *dSLC6A1* is expressed compared to wild-type controls. We also see significant differences when the *dSLC6A1^A334S^* is expressed compared to *dSLC6A1* and the UDN variant again performs as a hypomorph. These data emphasize the sensitivity of GABA transporter dysregulation, where insertion of the wild-type transgene induces significant phenotypes in regard to sleep and activity behavior. This data also highlights the extreme sensitivity of sleep latency. In this assessment of p.(A334S), the strongest phenotypes are seen in sleep consolidation, observed as a decrease in total sleep bout number and increase in sleep bout duration during the night. We also observe mild seizure sensitivity when *dSLC6A1^A334S^*is expressed. This in conjunction with the loss-of-function phenotypes observed when the p.(A334S) variant is expressed using a human *SLC6A1* transgene supports the conclusion that the UDN variant is a hypomorphic loss-of-function allele.

## Discussion

Even though *SLC6A1* is an established disease gene, variants of uncertain significance remain unresolved. Here we present the case of an individual identified through the UDN with a variant of uncertain significance in *SLC6A1.* Individuals with confirmed *SLC6A1*-related disorders were recruited to the BGR for clinical comparative analysis. The BGR initiative allows for comprehensive assessment of clinical phenotypes and the impact of variants in emerging neurodevelopmental genes including *SLC6A1*, to improve the diagnostic burden for rare disease cases^14^. The UDN also strives to improve the diagnostic rate for those living with rare disease through additional phenotypic and functional characterization. The UDN model organisms screening center allows for the evaluation of variant induced changes in protein function compared to the human reference protein. We perform clinical and functional genomic assessments to streamline and accelerate characterization of *SLC6A1*-related neurodevelopmental disorders.

In this study, we demonstrate the utility of deep clinical phenotyping in conjunction with functional analysis *in vivo* to resolve variants of uncertain significance. We leverage this approach to solve a case involving a variant in a known disease gene associated with variable expressivity. We identify an undiagnosed individual harboring an inherited p.(A334S) variant of uncertain significance presenting with cognitive disability, intractable seizures, and autism. Pedigree analysis reveals a variable clinical presentation among family members, with a sibling also harboring the variant who is only affected with autism, highlighting the complexity in diagnosis and treatment due to this variability. The p.(A334S) variant has not been observed in large population cohorts (gnomAD v4) and has a supporting *in silico* prediction (CADD Phred score v1.7 : 26.4) suggesting it is likely damaging. Additionally, another missense variant at this residue p.(A334P) has been associated with myoclonic-atonic seizures, further supporting the damaging nature of the p.(A334S) variant^33^. We identify a symptom profile for the UF1 that clusters similarly to other known cases of *SLC6A1*-related neurodevelopmental disorders.

We utilize HPO-based quantitative phenotypic clustering analysis to evaluate phenotypic subgroups as this approach is a promising method for generating new hypotheses regarding the role of *SLC6A1* variants in neurodevelopmental phenotypes. We find that the p.(A334S) variant clusters among other strong loss-of-function alleles and near another missense variant, p.(G297R) that is also associated with variable expressivity. This allele has been previously evaluated *in vivo* and displays significantly impaired GABA transport consistent with a strong loss-of-function mechanism^7^. The research presented here identifies that the UDN p.(A334S) variant and additional recurrent missense variants, p.(A288V) and p.(G297R) are associated with a variable presentation of symptoms including a mild to severe phenotypic spectrum. Our analysis also demonstrates that our frameshift variants cluster together with a more severe symptom profile including EEG abnormalities, tremors, and language impairment. These findings suggest that hypomorphic alleles, particularly missense variants, could be driving the clinical variability observed in *SLC6A1*-related neurodevelopmental disorders. While additional clinical research is needed to confirm this, the variability observed here strongly suggests the possibility that additional genetic or environmental factors are responsible for modulating *SLC6A1-*related disease.

Confirming the nature of candidate pathogenic variants is the first step toward targeted therapeutics. We evaluated the p.(A334S) variant *in vivo* in the fly model and found seizure, sleep, and reduced activity associated with *Gat* loss-of-function. We present evidence that the fly models generated as part of this study can be used to evaluate variant specific effects *in vivo*, and we identify sleep as a sensitive read out to capture partial loss-of-function phenotypes. In sleep studies, heterozygous loss of the fly ortholog *Gat* induces extremely reduced sleep latency, a result which is consistent with prior reporting of alterations in GABA signaling. We also found that co-expression of the human reference *SLC6A1* is sufficient to suppress this phenotype. Each of the variants tested in the human cDNA, including the UF1 variant fail to suppress sleep latency to wild-type levels, providing supportive evidence that all variants tested are putative hypomorphic alleles.

We also identified loss-of-function phenotypes when p.(A334S) is expressed in the context of the fly gene. We found seizure sensitivity, sleep, and reduced activity phenotypes are induced when a heterozygous variant transgene is expressed. These data identify that knock-in alleles are a promising approach to study dosage sensitive genes since we also identify reduced longevity when the human reference *SLC6A1* is over-expressed. The UF1 and additional individuals included in this study experience intractable epilepsy and GAT-1 targeted therapeutics could improve symptom management for affected individuals. Since we identify overexpression toxicity with the human *SLC6A1,* therapeutic approaches may need to modulate GAT-1 function in a narrow critical range. Future pre-clinical drug repurposing could identify therapeutics that are capable of suppressing seizures and improving behavioral phenotypes in these fly models. Identifying hypomorphic subgroups will be critical to support this research.

Additional studies could identify the effect of multiple loss-of-function variants in the GABA transporter/receptor pathway. This also highlights the utility of clinical exome or genome sequencing (ES/GS) in the diagnosis of *SLC6A1* neurodevelopmental disorders as compared to panel testing. The multigene panels for neurodevelopmental disorder may not include *SLC6A1* and other GABA transporter/receptor pathway genes, and the diagnostic sensitivity of the testing used for each gene vary by laboratory and are likely to change over time. Furthermore, additional variants in the GABA pathway that may be identified in the ES could prove to be promising therapeutic targets for improving symptoms.

We conclude that the UF1 variant p.(A334S) is a hypomorphic loss-of-function allele. Since we identify variable clinical presentation among related and unrelated individuals harboring the same variant, the results reported herein open up the possibility that *SLC6A1-*related neurodevelopmental disorders could be influenced by additional risk factors. It is important for a thorough genetic evaluation to be completed during clinical diagnosis to inform symptom progression and to identify additional genetic therapeutic targets to modulate seizure phenotypes. As the resolution improves for rare disease, it is likely that additional modifiers and risk factors will be identified. Thorough characterization of genetic and environmental factors will be critical in identifying the etiology of *SLC6A1* disorders. In all, comprehensive clinical and functional studies are essential to fully resolve variants of uncertain significance, even in genes associated with rare monogenic disorders.

## Data Availability

All data produced in the present work are contained in the manuscript

## Acknowledgements

We would like to thank the families for their involvement in this study. We are sincerely indebted to the generosity of the participants and their families in the Undiagnosed Diseases Network and Brain Gene Registry communities across the United States who contributed their time and effort to this study. We would also like to thank Amber Freed and the *SLC6A1 Connect* Foundation for their support of our research. We would like to acknowledge all members of the UDN model organisms screening center for their valuable input and insight. We would also like to thank Mei-Chu Huang for their help with molecular cloning techniques.

## Undiagnosed Diseases Network

Aaron Quinlan, Adeline Vanderver, Adriana Rebelo, Aimee Allworth, Alan H. Beggs, Alden Huang, Alex Paul, Ali Al-Beshri, Alistair Ward, Allyn McConkie-Rosell, Alyssa A. Tran, Andrea Gropman, Andrew B. Crouse, Andrew Stergachis, Anita Beck, Anna Hurst, Anna Raper, Anne Hing, Arjun Tarakad, Ashley Andrews, Ashley McMinn, Ashok Balasubramanyam, AudreyStephannie C. Maghiro, Barbara N. Pusey Swerdzewski, Ben Afzali, Ben Solomon, Beth A. Martin, Breanna Mitchell, Brendan C. Lanpher, Brendan H. Lee, Brent L. Fogel, Brett H. Graham, Brian Corner, Bruce Korf, Calum A. MacRae, Camilo Toro, Cara Skraban, Carlos A. Bacino, Carson A. Smith, Cecilia Esteves, Changrui Xiao, Chloe M. Reuter, Christina Lam, Christine M. Eng, Claire Henchcliffe, Colleen E. Wahl, Corrine K. Welt, Cynthia J. Tifft, Dana Kiley, Daniel Doherty, Daniel J. Rader, Daniel Wegner, Danny Miller, Daryl A. Scott, Dave Viskochil, David A. Sweetser, David R. Adams, Dawn Earl, Deborah Barbouth, Deborah Krakow, Deepak A. Rao, Devin Oglesbee, Devon Bonner, Donna Novacic, Dustin Baldridge, Edward Behrens, Edwin K. Silverman, Elaine Seto, Elijah Kravets, Elizabeth A. Burke, Elizabeth Blue, Elizabeth L. Fieg, Elizabeth Rosenthal, Ellen F. Macnamara, Elsa Balton, Emilie D. Douine, Emily Glanton, Emily Shelkowitz, Eric Allenspach, Eric Klee, Eric Vilain, Erin Baldwin, Erin Conboy, Erin E. Baldwin, Erin McRoy, Esteban C. Dell’Angelica, Euan A. Ashley, F. Sessions Cole, Filippo Pinto e Vairo, Frances High, Francesco Vetrini, Francis Rossignol, Fuki M. Hisama, Gabor Marth, Gail P. Jarvik, Gary D. Clark, George Carvalho, Gerard T. Berry, Ghayda Mirzaa, Gill Bejerano, Giorgio Sirugo, Gonench Kilich, Guney Bademci, Heidi Wood, Herman Taylor, Holly K. Tabor, Hongzheng Dai, Hsiao-Tuan Chao, Hugo J. Bellen, Ian Glass, Ian R. Lanza, Ingrid A. Holm, Isaac S. Kohane, Ivan Chinn, J. Carl Pallais, Jacinda B. Sampson, James P. Orengo, Jason Hom, Jennefer N. Kohler, Jennifer E. Posey, Jennifer Wambach, Jessica Douglas, Jiayu Fu, Jill A. Rosenfeld, Jimann Shin, Jimmy Bennett, Joan M. Stoler, Joanna M. Gonzalez, John A. Phillips III, John Carey, John J. Mulvihill, Joie Davis, Jonathan A. Bernstein, Jordan Whitlock, Jose Abdenur, Joseph Loscalzo, Joy D. Cogan, Julian A. Martínez-Agosto, Justin Alvey, Kahlen Darr, Kaitlin Callaway, Kathleen A. Leppig, Kathleen Sullivan, Kathy Sisco, Kathyrn Singh, Katrina Dipple, Kayla M. Treat, Kelly Hassey, Kelly Schoch, Kevin S. Smith, Khurram Liaqat, Kim Worley, Kimberly Ezell, Kimberly LeBlanc, Kumarie Latchman, Lance H. Rodan, Laura Pace, Laurel A. Cobban, Lauren C. Briere, Leoyklang Petcharet, LéShon Peart, Lili Mantcheva, Lilianna Solnica-Krezel, Lindsay C. Burrage, Lindsay Mulvihill, Lisa Schimmenti, Lisa T. Emrick, Lorenzo Botto, Lorraine Potocki, Lynette Rives, Lynne A. Wolfe, Manish J. Butte, Margaret Delgado, Maria T. Acosta, Marie Morimoto, Mariko Nakano-Okuno, Mark Wener, Marla Sabaii, Martha Horike-Pyne, Martin G. Martin, Martin Rodriguez, Matt Velinder, Matthew Coggins, Matthew Might, Matthew T. Wheeler, Maura Ruzhnikov, MayChristine V. Malicdan, Meghan C. Halley, Melissa Walker, Michael Bamshad, Michael F. Wangler, Miguel Almalvez, Mohamad Mikati, Monika Weisz Hubshman, Monte Westerfield, Mustafa Tekin, Naghmeh Dorrani, Neil H. Parker, Neil Hanchard, Nicholas Borja, Nicola Longo, Nicole M. Walley, Nina Movsesyan, Nitsuh K. Dargie, Oguz Kanca, Orpa Jean-Marie, Page C. Goddard, Paolo Moretti, Patricia A. Ward, Patricia Dickson, Paul G. Fisher, Pengfei Liu, Peter Byers, Pinar Bayrak-Toydemir, Precilla D’Souza, Queenie Tan, Rachel A. Ungar, Rachel Mahoney, Ramakrishnan Rajagopalan, Raquel L. Alvarez, Rebecca C. Spillmann, Rebecca Ganetzky, Rebecca Overbury, Rebekah Barrick, Richard A. Lewis, Richard L. Maas, Rizwan Hamid, Rong Mao, Ronit Marom, Rosario I. Corona, Russell Butterfield, Sam Sheppeard, Sanaz Attaripour, Seema R. Lalani, Serena Neumann, Shamika Ketkar, Shamil R. Sunyaev, Shilpa N. Kobren, Shinya Yamamoto, Shirley Sutton, Shruti Marwaha, Sirisak Chanprasert, Stanley F. Nelson, Stephan Zuchner, Stephanie Bivona, Stephanie M. Ware, Stephen Pak, Steven Boyden, Suman Jayadev, Surendra Dasari, Susan Korrick, Suzanne Sandmeyer, Tahseen Mozaffar, Tammi Skelton, Tara Wenger, Terra R. Coakley, Thomas Cassini, Thomas J. Nicholas, Timothy Schedl, Tiphanie P. Vogel, Vaidehi Jobanputra, Valerie V. Maduro, Vandana Shashi, Virginia Sybert, Vishnu Cuddapah, Wendy Introne, Wendy Raskind, Willa Thorson, William A. Gahl, William E. Byrd, William J. Craigen, Yan Huang, Yigit Karasozen,

## Brain Gene Registry Consortium

**Principal Investigators:** Wasserstein, M^1^; Chopra, M^2^; Sahin, M ^2^; Wangler, M^3^; Schultz, B^4^; Izumi, K^4^; Gropman, A^5^; Smith-Hicks, C^6^; Abbeduto, L^7^; Hazlett, H^8^; German, K^9^; DaWalt, L^10^; Neul, J^11^; Constantino, J^12^; Payne, PRO^13^

**Co-Investigators:** Gurnett, C^13^; Baldridge D^13^; Srivastava, S^2^; Molholm, S^1^; Walkley, S^1^; Storch, E^3^; Samaco, R^3^; Cohen, J^6^; Shankar, S^7^; Piven, J^8^, Berger, S^5^

**Clinical Coordinating Center:** Mahida, S^2^; Sveden, A^2^; Dies, K^2^

**ClinGen Genome Connect Team:** Riggs, ER^14^; Savatt, JM^14^

**Program Management and CIELO Team:** Lanzotti, V ^13^; Oh, I^13^; Gupta, A^13^; Minor, B^13^

## Affiliations

1. Albert Einstein College of Medicine
2. Boston Children’s Hospital
3. Baylor College of Medicine
4. Children’s Hospital of Philadelphia
5. Children’s National Medical Center
6. Kennedy Krieger Institute
7. UC Davis MIND institute
8. University of North Carolina, Chapel Hill
9. University of Washington
10. Waisman Center, University of Wisconsin-Madison
11. Vanderbilt University Medical Center
12. Pediatric Institute, Emory University School of Medicine and Children’s Healthcare of Atlanta
13. Washington University School of Medicine in St. Louis
14. Autism and Developmental Medicine Institute, Geisinger, Danville PA

## Ethics declaration

Probands were recruited through their local referring physicians and the UDN clinical site at Vanderbilt University. UF1 was identified through the Undiagnosed Diseases Network (UDN), and individuals BGR Family 1-13 were identified through the National Brain Gene Registry. Prior to inclusion, informed written consent was obtained from the legal guardians of the individuals included in this study for research and publication according to the standards and practices of the institutional review board and ethics committee at Vanderbilt University. Documents and consent forms were standardized according to the requirements of the UDN.

## Supplementary Figures and Table

**Figure S1** Phenotypic depth. The number of HPO terms per individual is plotted. On average, there are 12 HPO terms per individual noted for further analysis.

**Figure S2** RNAP domain scores. Seven RNAP domain scores (%) are plotted along with percentage of answered questions for each individual per domain. A greater domain score indicates a worse phenotype noted in the individual.

**Figure S3** Annotation grid of BGR8_p.G297R. This is a case of dual diagnosis with variants in both *SLC6A1* and *PTPN11*, both explain the totality of the phenotypes noted in the individual.

**Figure S4** Human *SLC6A1* suppresses bang sensitivity in flies. (A). Expression of *SLC6A1^Ref^, SLC6A1^A288V^, SLC6A1^S295^L, SLC6A1^G297R^,* or *SLC6A1^A334S^* with *Gat^TG4^* does not result in seizure sensitivity. (B) Homozygous loss of *Gat* (*Gat^TG4^ / Df(4)Ed6381*) induces bang sensitivity that is rescued by co-expression of *SLC6A1^Ref^.* (C) Loss of *Gat* results in reduced longevity, and expression of *SLC6A1^Ref^* with *Gat^TG4^* induces a more severe reduced longevity phenotype.

**Figure S5** Day and night sleep are more fragmented when *SLC6A1^A334S^* is expressed. (A) Total day sleep is not affected but total night sleep is decreased for *SLC6A1^A288V^* (P=0.011), *SLC6A1^G297R^* (P<0.0001), *SLC6A1^A334S^* (P<0.0001). (B) Sleep bout number is increased during the day for *SLC6A1^A288V^* (P<0.0001), *SLC6A1^S295L^* (P<0.0001), *SLC6A1^G297R^* (P<0.0001), *SLC6A1^A334S^* (P<0.0001) and night *SLC6A1^A288V^* (P<0.0001), *SLC6A1^S295L^* (P<0.0001), *SLC6A1^G297R^* (P<0.0001), and *SLC6A1^A334S^* (P<0.0001). (C) Sleep bout length is decreased during the day for *SLC6A1^A288V^* (P<0.0001), *SLC6A1^S295L^* (P<0.0001), *SLC6A1^G297R^* (P<0.0001), and *SLC6A1^A334S^* (P<0.0001) and night *SLC6A1^A288V^* (P<0.0001), *SLC6A1^S295L^* (P<0.0001), *SLC6A1^G297R^* (P<0.0001), and *SLC6A1^A334S^* (P<0.0001). (D) Total activity is increased during the day for *SLC6A1^S295L^*(P=0.0014) and increased during the night for *SLC6A1^A288V^* (P=0.0001), *SLC6A1^G297R^* (P<0.0001), *SLC6A1^A334S^* (P<0.0001). Welch’s ANOVA with Dunnett’s T3 multiple comparisons test.

**Figure S6** Expression of the UDN variant promotes night sleep consolidation. (A) There is a decrease in *dSLC6A1^A334S^* total day sleep (P<0.0001) and an increase in total night sleep (P<0.0001) compared to *dSLC6A1*. (B) There is an increase in *dSLC6A1^A334S^*day sleep bout number (P<0.0001) and no significant change in night bout number. (C) There is a decrease in *dSLC6A1^A334S^*day sleep bout length (P=0.0006) and an increase in night bout length (P=0.0254). (D) There is an increase in *dSLC6A1^A334S^* total day activity (P<0.0001) and a decrease in total night activity *dSLC6A1^A334S^* (P<0.001). Welch’s ANOVA with Dunnett’s T3 multiple comparisons test.

**Table S1** Genotype and phenotypes of individuals with *SLC6A1* variants as noted in this study.

