## Supplementary material for "Resolution of *SLC6A1* variable expressivity in a multi-generational family using deep clinical phenotyping and *Drosophila* models": Figure S1

Supplemental Figures

Figure S1

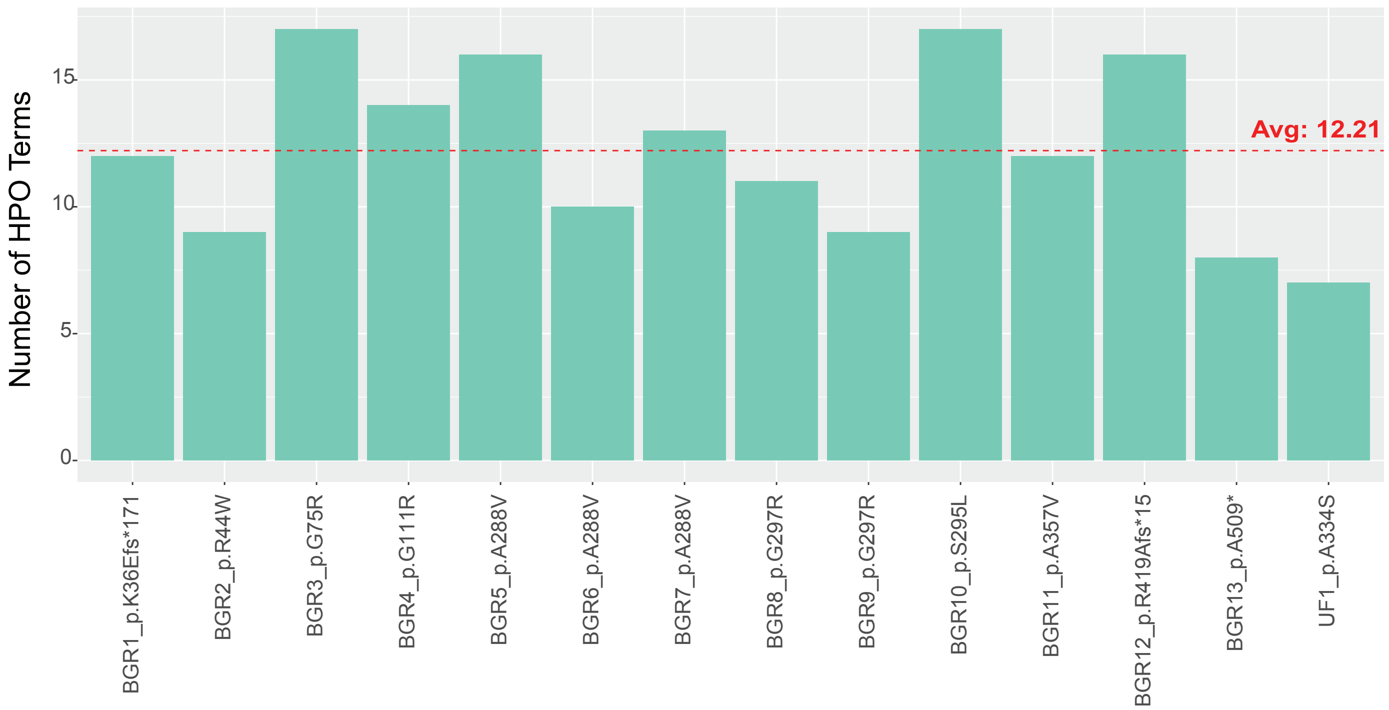


Figure S1: Phenotypic depth. The number of HPO terms per individual is plotted. On average, there are 12 HPO terms per individual noted for further analysis.
