## Supplementary material for "Resolution of *SLC6A1* variable expressivity in a multi-generational family using deep clinical phenotyping and *Drosophila* models": Figure S2

### Supplemental Figures

#### Figure S2

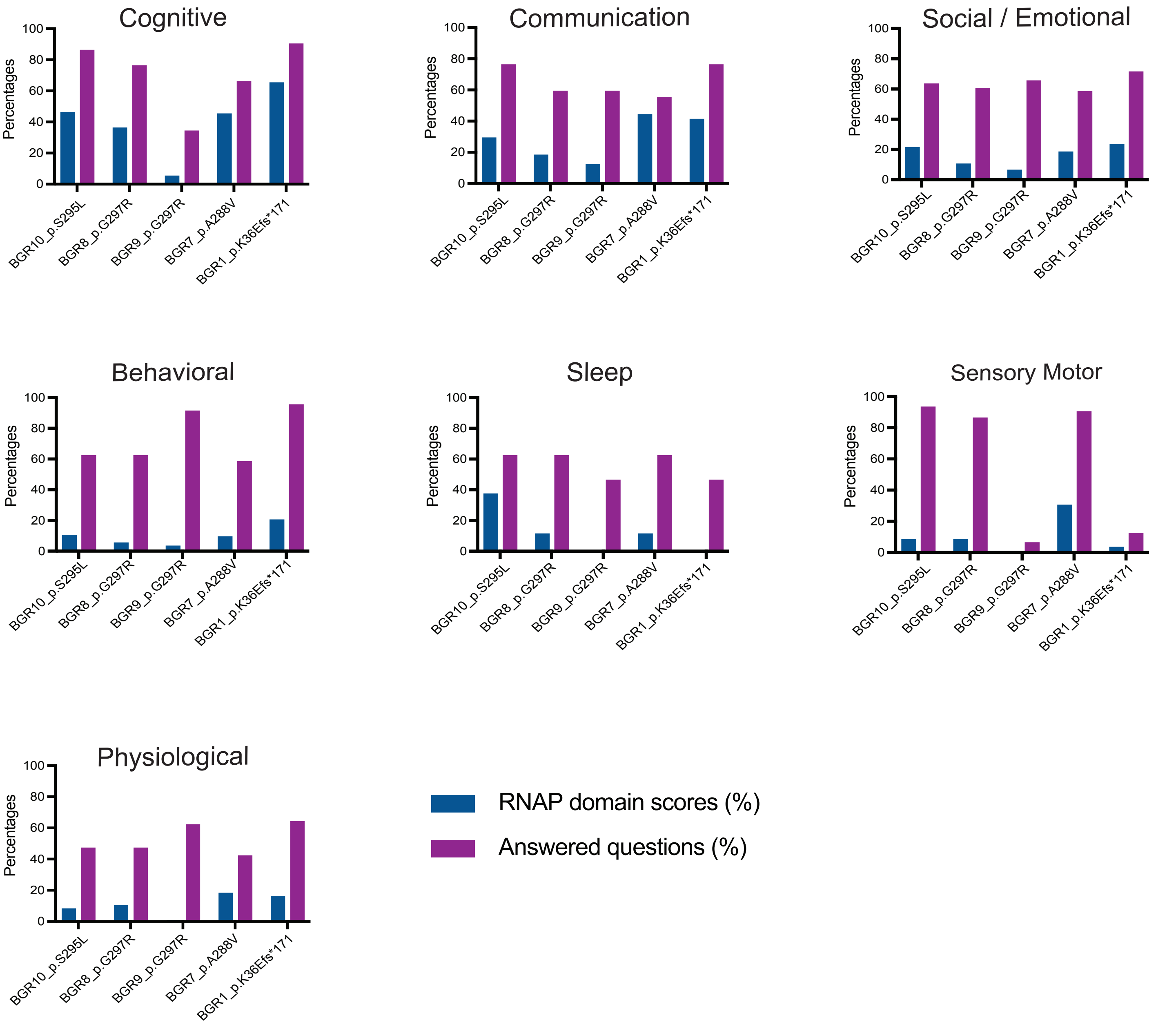


Figure S2: RNAP domain scores. Seven RNAP domain scores (%) are plotted along with percentage of answered questions for each individual per domain. Greater the percentage of domain score wrose the phenotype noted in the individual.
