## Supplementary material for "Resolution of *SLC6A1* variable expressivity in a multi-generational family using deep clinical phenotyping and *Drosophila* models": Figure S3

Supplemental Figures

Figure S3

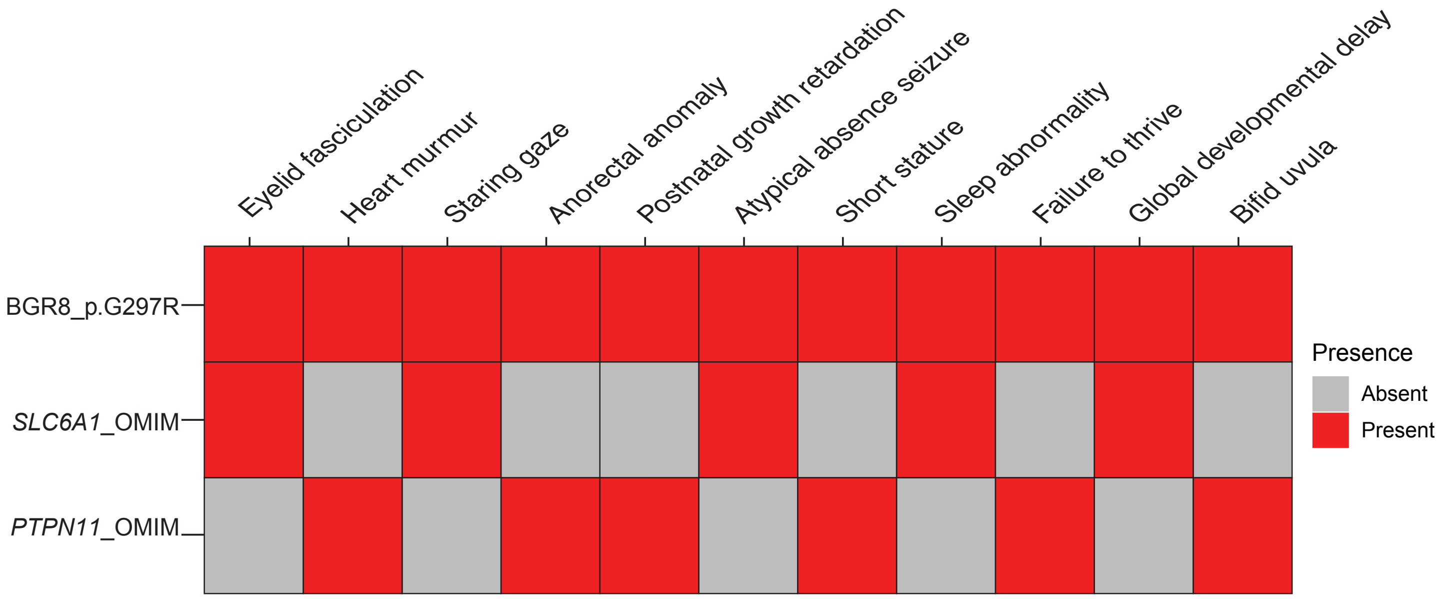


Figure S3: Annotation grid of BGR_Family08_p.G297R. This is a case of dual diagnosis with variants in both *SLC6A1* and *PTPN11*, both explain the totality of the phenotypes noted in the individual.
