## Supplementary figures and images for "Resolution of *SLC6A1* variable expressivity in a multi-generational family using deep clinical phenotyping and *Drosophila* models"

### Figure S4

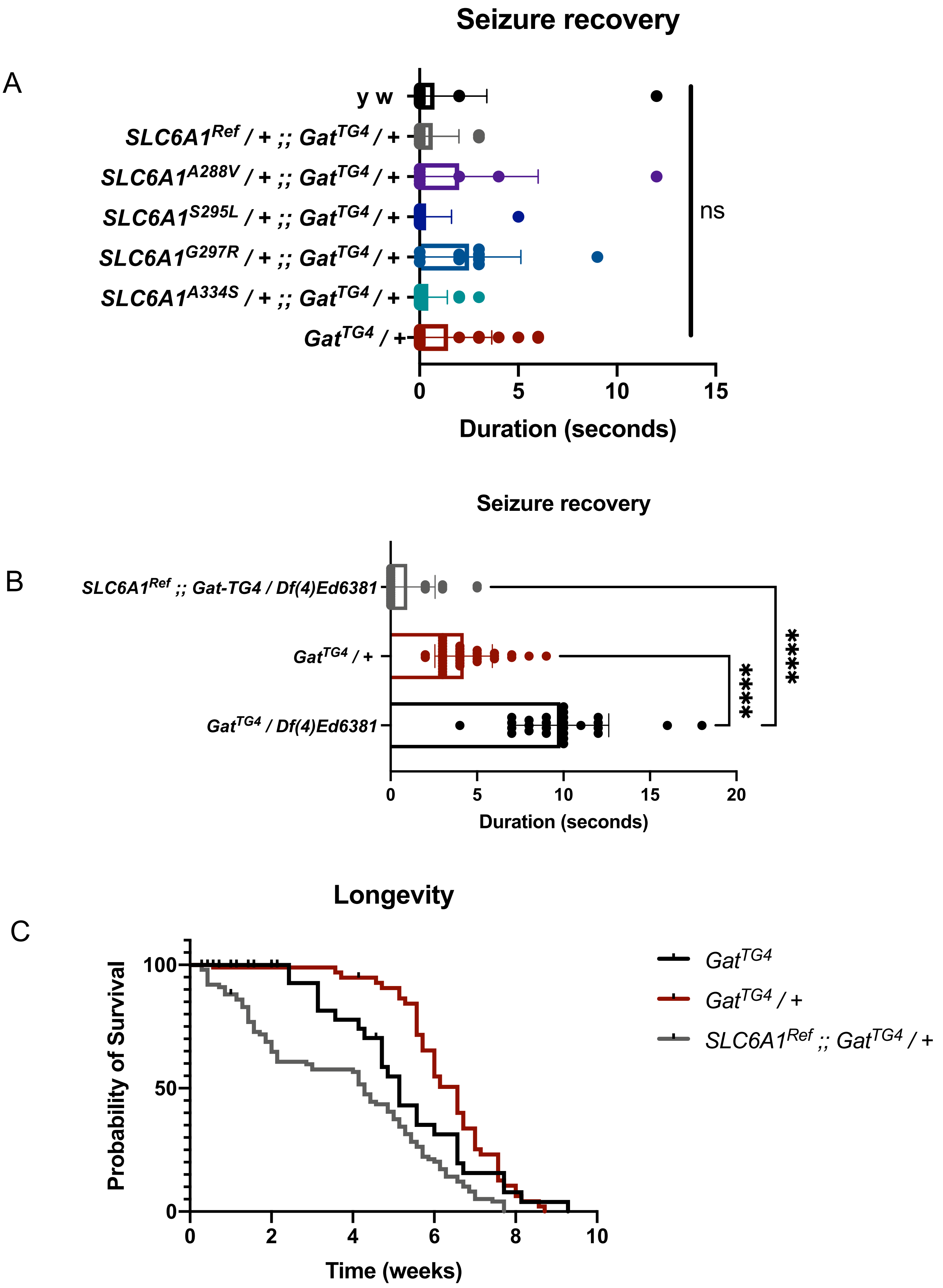

### Figure S5

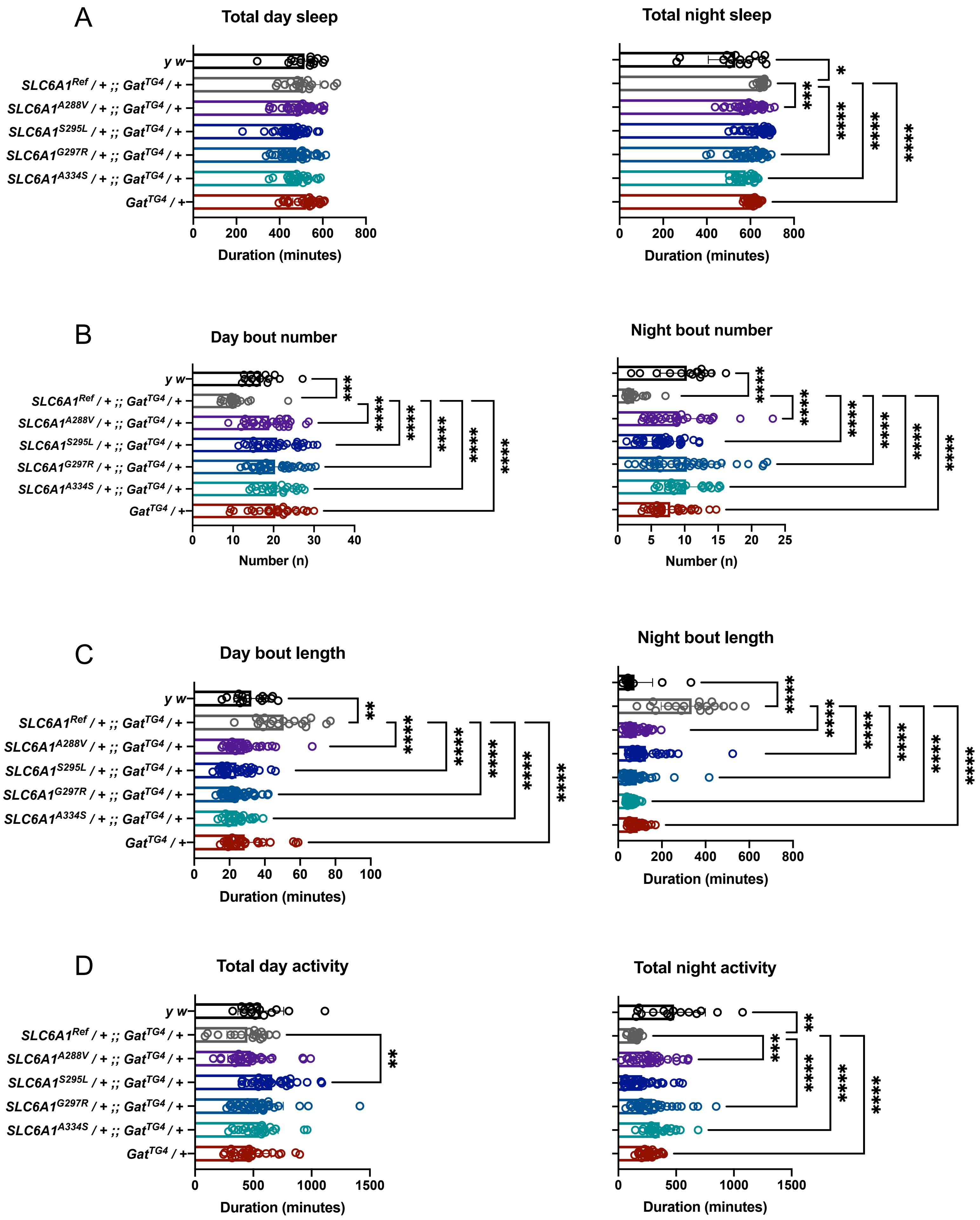

### Figure S6

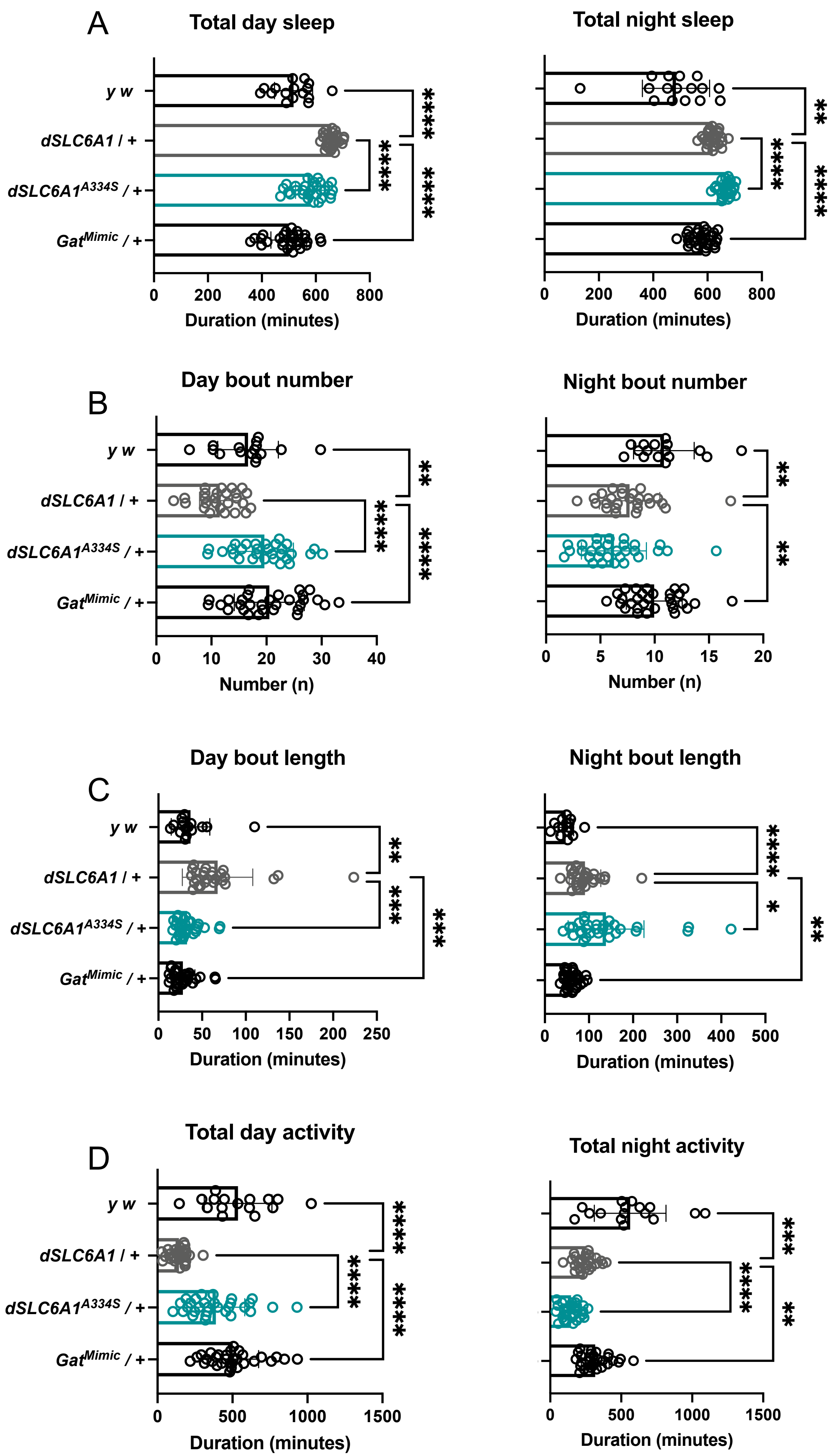
